# A meta-analysis of 20 years of data on people who inject drugs in metropolitan Chicago to inform computational modeling

**DOI:** 10.1101/2021.02.24.21252385

**Authors:** Basmattee Boodram, Mary Ellen Mackesy-Amiti, Aditya Khanna, Bryan Brickman, Harel Dahari, Jonathan Ozik

## Abstract

Progress toward hepatitis C virus (HCV) elimination in the United States is not on track to meet targets, as injection drug use continues to drive increasing HCV incidence. Computational models are useful in understanding the complex interplay of individual, social, and structural level factors that might alter HCV incidence, prevalence, transmission, and disease progression. However, these models need to be informed with real socio-behavioral data. We conducted a meta-analysis of research studies spanning 20 years of research and interventions with people who inject drugs in metropolitan Chicago to produce parameters for a synthetic population for a computational model. We then fit an exponential random graph model (ERGM) using the network estimates from the meta-analysis in order to develop the dynamic component of a realistic agent-based model.

## Introduction

Hepatitis C virus (HCV) infection is a leading cause of chronic liver disease and mortality worldwide. An estimated 71.1 million are chronically infected globally [1]. Persons who inject drugs (PWID) are at the highest risk for acquiring and transmitting HCV infection primarily through syringe-sharing [2]. Despite the long-term availability of harm reduction strategies such as syringe services programs (SSPs), medication-assisted therapy (MAT), behavioral counseling and recent availability of highly-effective direct-acting antiviral (DAA) treatment, injection drug use (IDU) and HCV incidence have been increasing at alarming rates among PWID in the United States. The U.S. lags behind other high-income countries in progress toward HCV elimination goals [3]. To achieve the World Health Organization’s (WHO) goal of reducing new chronic infections by 90% and mortality by 65% by 2030 [4], we must better understand the heterogeneous PWID sub-populations who may be differentially driving incidence, prevalence, and/or liver disease progression. For example, prior studies have shown that HCV incidence is highest among young PWID, while mortality is highest among African Americans [5]. Elimination of HCV will require combating both HCV transmission and low DAA treatment initiation and completion as well as understanding the complex interplay of individual, social, and structural level factors that might alter HCV incidence, prevalence, transmission, and disease progression through specific combinations of prevention, intervention and treatment strategies. This level of complexity cannot be addressed with traditional empirical studies and is best suited for sophisticated computational models.

Our study reports on the results of a meta-analysis of recent studies (1997-2017) [6-16] among PWID to better characterize this population using data representing the estimated 32,000 PWID who reside in metropolitan Chicago, Illinois [17]. The synthesized data will be used to 1) better inform interventions, and 2) to provide a realistic profile of a synthetic population for a computational model that could be used to examine specific combinations of HCV elimination strategies. We also fit an exponential random graph model (ERGM) using the network estimates from the meta-analysis in order to develop the dynamic component of a realistic agent-based model (ABM). ERGMs offer a uniquely robust and flexible approach in modeling the dependencies between behavioral parameters, and have been used to model syringe sharing networks in a number of related contexts [18].

## Methods

### Meta-analysis Data Sources

Twelve data sources were included in the meta-analysis, including eleven research studies and one syringe service program (SSP) (Table 1). We selected these diverse datasets on the basis that they would provide rich sources of data for developing the highly heterogeneous PWID synthetic population for the model. SSP data were split into two datasets according to year of enrollment to represent earlier and later time periods. In all datasets, records only included individuals 18 years or older and excluded those missing key demographic information (sex, age, race/ethnicity). Subjects who identified as transgender were excluded if the study did not include a variable to classify them as male or female (i.e. biological sex). The datasets were stratified by sex (male or female), race/ethnicity (non-Hispanic White, non-Hispanic Black, Hispanic, and Other/mixed), and age category (18-24, 25-34, 35-44, 45 and older). Measures were harmonized across studies, and frequencies of categorical variables and means and standard errors of continuous variables were computed for each strata in each dataset. Poisson standard errors were computed for count variables such as network size and number of sex partners.

**Table 1.** Data sets included in the PWID meta-analysis.

| Study | Years | Sample size | Age range | Reference |
| --- | --- | --- | --- | --- |
| CIDUS 2 | 1997-1999 | 695 | 18-34 | [6] |
| CIDUS 3 | 2002-2004 | 775 | 18-34 | [7] |
| NATHCV | 2002-2005 | 110 | 18-34 | [8] |
| NIHU | 2004-2006 | 150 | 18-34 | [9] |
| SATHCAP | 2005-2009 | 1356 | 18 + | [10] |
| PSYCH | 2008-2010 | 610 | 18-25 | [11] |
| NHBS IDU2 | 2009 | 553 | 18 + | [12] |
| NHBS IDU3 | 2012 | 210 | 18 + | [13] |
| SOCNET | 2012-2013 | 164 | 18-34 | [14] |
| NHBS IDU4 | 2015 | 541 | 18 + | [15] |
| MOOD | 2016-2017 | 180 | 18-34 | [16] |
| COIP SEP (1) | 2005-2010 | 4952 | 18 + |  |
| COIP SEP (2) | 2011-2016 | 2632 | 18 + |  |

PWID: People who inject drugs
CIDUS: Collaborative Injection Drug Users Study (CDC)
NATHCV: Early natural history of HCV infection among injection drug users (NIDA)
NIHU: Non-injected heroin use, HIV, and transitions to injection (NIDA)
SATHCAP: Sexual Acquisition and Transmission of HIV Cooperative Agreement Project (NIDA)
PSYCH: Psychiatric disorders and HIV risk behaviors among young injection drug users (NIDA)
SOCNET: Social networks of young injection drug users (Chicago DCFAR)
MOOD: Emotion dysregulation and risky behavior among people who inject drugs (NIDA)
NHBS: National HIV Behavioral Surveillance (CDC)

### Measures

#### Sociodemographic characteristics

Measures included basic demographic categories of sex, age, race/ethnicity, and place of residence (dichotomized as within the city limits of Chicago (urban) vs. all other surrounding areas). Measures of current employment, income, and recent homelessness were also available. Several studies asked about sources of income, while others asked directly about employment status. The measures were harmonized by creating indicators of any employment (including temporary work), and regular employment (full or part-time). Measures of income were answered on a categorical scale, in increments of $500 or $5000. We used the median of the category (the lower limit for the top category) as the value of income. Homelessness was measured as self-perceived homelessness (e.g. have you been homeless, have you considered yourself homeless) within the past 6 or 12 months.

#### Harm reduction

We included measures of current substance use treatment (other than peer support groups), and obtaining syringes from a syringe service program (SSP) in recent months.

#### IDU networks

Two measures of IDU network size were available: 1) number of people respondent knows who inject drugs (total PWID network, 6 studies); 2) number of people respondent injected drugs with in the past 30 days, 3 months, or 6 months (injection sub-network, 5 studies). Three studies included both of these measures. The syringe-sharing network is a further subset of the network of people injected with. We estimated the number of people respondent used a syringe after (in-degree), and the number of people respondent gave his or her syringe to after using it (out-degree). As measures of network mixing, we estimated the network proportions by sex, race/ethnicity, and Chicago/non-Chicago residence, based on responses to questions included in studies that used respondent driven sampling (RDS). However, only one study (SATHCAP) included respondents of all ages, therefore for purposes of informing the model, network mixing estimates were based on this study only.

#### IDU behaviors

Variables related to injection drug use included age of first injection drug use, years of injecting (computed from current age and age of first injection), and frequency of injection in past 30 days. Frequency of injection was estimated based on responses to two questions: 1) average frequency of injection in the past 3, 6 or 12 months, with categorical responses, from once a month or less to every day, and 2) typical number of times injected per day, from once to 10 or more times a day.

Injection risk behaviors included syringe-mediated drug sharing (SMS) in the past 3, 6, or 12 months (injected drugs with a syringe after someone else squirted drugs into it from another syringe, “backloading”, or drugs were mixed, measured, or divided using someone else’s syringe), any receptive syringe sharing (RSS; injected with a syringe that someone else had used to inject) in the past 30 days to 12 months, frequency of RSS (estimated percent of injections that involved RSS based on Likert scale response or number of RSS injections and frequency of injection), any equipment sharing (shared cookers, cotton, or rinse water) in the past 30 days to 12 months, frequency of sharing cookers (converted Likert scale responses to percentage of injections).

### Meta-analysis: Random Effects Model

Under the fixed-effect model it is assumed that the true effect size for all studies is identical, and the only reason the effect size varies between studies is due to sampling error. By contrast, under the random-effects model the goal is not to estimate one true effect, but to estimate the mean of a distribution of effects [19]. Although our studies have many similarities, there may be important differences between the study populations that influence the observed effect sizes, including most obviously time. There is also variability in the measures used across the studies that can affect effect sizes, e.g. reporting on behavior in the past 3 months vs. past 6 months.

Estimates of network size and injection risk behavior were stratified by sex, race and ethnicity (non-Hispanic white, non-Hispanic Black, Hispanic (all races), and non-Hispanic other races), and age category (18-24, 25-34, 35-44, and 45 and older.) Meta-analysis of proportions was conducted using the -*metaprop*-command in Stata [20]. The Freeman-Tukey double arcsine transformation [21] was employed to stabilize the variances [22], and exact confidence intervals were computed. Meta-analysis of continuous and count variables was conducted using the -*metan*-command in Stata [23] to estimate a random effects model using the method of DerSimonian and Laird [24]. Poisson standard errors were used in the estimation of count variables. Sample estimates based on subgroups of fewer than five subjects were excluded.

### Mixed Effects Regression Analyses

We estimated mixed effects regression models in Stata (v. 15, StataCorp) to examine main effects of demographic variables on socioeconomic status, harm reduction, networks, and risk behavior. Logistic regression models were estimated for binary outcomes, and negative binomial regression models were estimated for count outcomes. Contrasts were computed to test the joint effects of multi-category variables of race/ethnicity and age category.

### Modeling Networks of Persons Who Inject Drugs

The theoretical framework for modeling IDU networks is based upon the exponential random graph models (ERGMs) [25-28]. The ERGMs are fit using the *ergm* package [29] in the R programming language [30] to generate directed graphs that represent the syringe sharing network. Conceptually, ERGMs can be understood to be similar to logistic regression models, where the outcome variable indicates presence and absence of an “edge” (also known as “tie” or “relational information”) and the independent variables describe network configurations.

Because PWID syringe-sharing relationships are not random[31], it is necessary to include mixing parameters in the model that govern the probability of tie formation. Characteristics such as sex, age, race and ethnicity, and geographic proximity are important factors that influence both tie formation and risk behaviors related to HCV transmission and acquisition [31-34].

The log-odds of formation of each partnership type were dependent upon the number and distribution of existing syringe-sharing relationships (henceforth, “relationships”) within the network. The mean number of such relationships was estimated from meta-analyses as described above, based on reported numbers of receptive and distributive syringe sharing partners, and the distribution of relationships was determined by the following parameters: mixing based on sex (“male” and “female”), race/ethnicity (“non-Hispanic white”, “non-Hispanic Black”, “Hispanic”, “non-Hispanic other”) and age (“under 25” vs. “25 and older”); the distribution of geographic distances and the distribution of in- and out-degree edges. The mathematical formulation of the model is given in Equation A.1 in the Appendix (S1 File). Standard tools in the *statnet* package were used to assess model fit and convergence. One hundred networks were simulated from the fitted model and the distributions of the simulated parameters were compared to the targets estimated from the meta-analysis above.

## Results

### Meta-analysis estimates

#### Socioeconomic status

On average 45% of PWID reported having some kind of employment including temporary work, and 26% of PWID were employed in a regular job (full or part-time). Women were less likely to be employed in any capacity (OR = 0.49, 95% CI 0.36 – 0.65), as were Hispanic (vs. non-Hispanic white, OR = 0.67, 95% CI 0.54 – 0.84) and non-Hispanic Black persons (vs. non-Hispanic white, OR = 0.63, 95% CI 0.42 – 0.93), and persons over 25 years old (Chi2[1] = 48.25, p<.0001). Results were similar for regular employment except that Chicago residence decreased the likelihood of regular employment (OR = 0.76, 95% CI 0.61 – 0.94). The average monthly income was $1,369, with higher income reported by non-Hispanic white PWID (Chi2[1] = 12.84, p = .0003), and decreasing with increasing age (Chi2[3] = 126.91, p<.0001). An estimated 39% of PWID reported being homeless; those who resided in Chicago were more likely to report being homeless (OR = 1.42, 95% CI 1.16 – 1.75), while non-Hispanic Black PWID were less likely to be homeless (35% vs. 44% non-Hispanic white, OR = 0.68, 95% CI 0.56 – 0.83).

#### Injection behavior and harm reduction

The estimated average age of first injection was 22 (95% CI 20.9 – 23.3), duration of injection was 9.9 years (95% CI 7.5 – 12.3), and average frequency of injection in the past 30 days was 75 times (95% CI 63.3 – 86.5). Over half of PWID (58%) reported use of a SSP, and 13% reported current substance use treatment. Women (OR = 1.23, 95% CI 1.04 – 1.46) and PWID over 25 (Chi2[1] = 10.07, p = .0015) were more likely to use a SSP, while non-Hispanic Black PWID were less likely than others to use a SSP after adjusting for age (Chi2[3]=22.80, p<.0001). PWID who resided in Chicago were marginally more likely to report using a SSP (OR = 1.28, 95% CI 1.00 – 1.64). Women were more likely than men to report current treatment (OR = 1.42, 95% CI 1.25 – 1.61), and current treatment increased with increasing age (Chi2[3] = 424.21, p<.0001).

#### Network size

Estimates of total PWID network and injection sub-network are shown in Tables 2 and 3. Total PWID network size tended to increase with age (Wald chi2[3] = 31.71, p<.0001), and there were no consistent sex or race/ethnicity differences. Injection sub-network size tended to be smaller in the oldest (45+) age category (chi2[1]=6.35, p=.01), and non-Hispanic white PWID tended to report a larger number of people they injected with compared to other categories (chi2[1]=12.28, p=.0005).

**Table 2.** Meta-analysis estimates of total PWID network size, by sex, race or ethnicity, and age category.

| Sex | Age category | non-Hispanic white |  |  | non-Hispanic Black |  |  | Hispanic |  |  | Other non-Hispanic |  |  |
| --- | --- | --- | --- | --- | --- | --- | --- | --- | --- | --- | --- | --- | --- |
|  |  | Mean | LL | UL | Mean | LL | UL | Mean | LL | UL | Mean | LL | UL |
| Male | 18-25 | 9.66 | 7.72 | 11.60 | NA |  |  | 7.25 | 6.12 | 8.38 | 7.33 | 6.18 | 8.49 |
|  | 25-34 | 15.77 | 7.38 | 24.15 | 13.44 | 11.97 | 14.90 | 15.25 | 9.99 | 20.50 | 4.39 | 3.20 | 5.57 |
|  | 35-44 | 22.18 | 15.76 | 28.61 | 22.52 | 11.44 | 33.60 | 21.66 | 16.32 | 27.00 | NA |  |  |
|  | 45+ | 20.81 | 17.46 | 24.16 | 22.22 | 19.87 | 24.58 | 28.03 | 17.94 | 38.13 | 16.64 | 9.64 | 23.64 |
| Female | 18-25 | 10.67 | 7.70 | 13.63 | NA |  |  | 5.85 | 3.09 | 8.61 | 8.12 | 6.76 | 9.47 |
|  | 25-34 | 16.53 | 8.40 | 24.65 | NA |  |  | 17.50 | 10.01 | 25.00 | NA |  |  |
|  | 35-44 | 23.18 | 15.90 | 30.45 | 17.62 | 6.15 | 29.09 | 23.94 | 17.00 | 30.88 | NA |  |  |
|  | 45+ | 20.85 | 16.68 | 25.01 | 21.76 | 12.15 | 31.37 | 28.10 | 17.62 | 38.58 | NA |  |  |

LL: lower limit 95% confidence interval; UL upper limit 95% confidence interval; NA: estimate not available, insufficient data

**Table 3.**
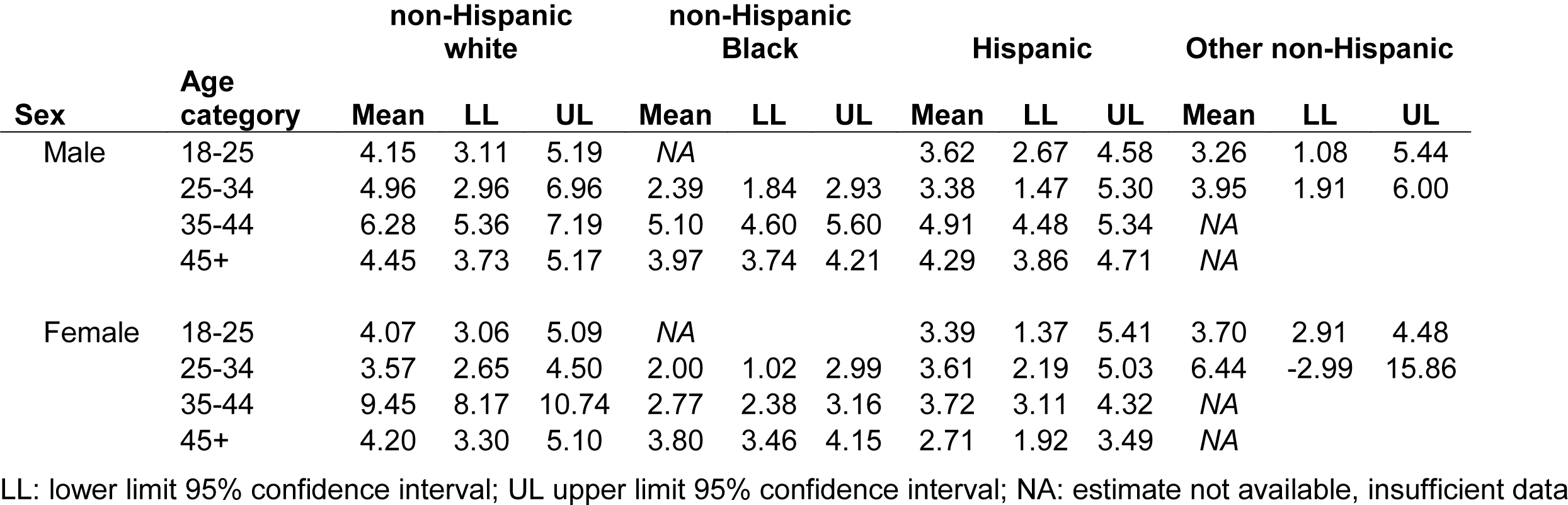
Meta-analysis estimates of injection drug use network size, by sex, race or ethnicity, and age category.

#### Network mixing

On average, men and women reported respectively that 69% (95% CI 0.67 – 0.70) and 61% (95% CI 0.58 – 0.64) of the people they knew who injected drugs were male. Young PWID (under 25 years old) reported on average 61% (95% CI 0.46 – 0.76) of their PWID network was also under 25, while older PWID reported 14% (95% CI 0.13 – 0.15) of their PWID network was under 25. Among PWID who lived in the city of Chicago, 87% (95% CI 0.82 – 0.93) of network members also lived in Chicago, while among PWID who lived in surrounding suburban areas or nearby states, 37% (95% CI 0.20 – 0.54) of network members lived in Chicago. Non-Hispanic white PWID reported that 74% (95% CI 0.61 – 0.87) of their network members were also white, non-Hispanic Black PWID reported that 55% of their network members were also non-Hispanic Black, and Hispanic PWID reported that 51% (95% CI 0.36 – 0.66) of their network members were also Hispanic.

#### Injection risk behavior

Overall, the estimated proportion of equipment sharing was 0.62 (95% CI 0.55-0.70), receptive syringe sharing (RSS) was 0.31 (95% CI 0.24-0.40), distributive syringe sharing (DSS) was 0.40 (95% CI 0.34-0.46), and syringe mediated sharing was 0.25 (95% CI 0.21 – 0.29). The average proportion of injections involving RSS was 0.12 (95% CI 0.09 – 0.15), and the average proportion of injections involving shared equipment was 0.28 (95% CI 0.23 – 0.33). The results for proportions of PWID in each demographic subgroup reporting any receptive and distributive syringe sharing are shown in Figs 1 and 2. Additional results are available in online supplemental materials (S2 File). Of note, several subgroups were small leading to large confidence intervals for the estimates.

**Fig 1.**
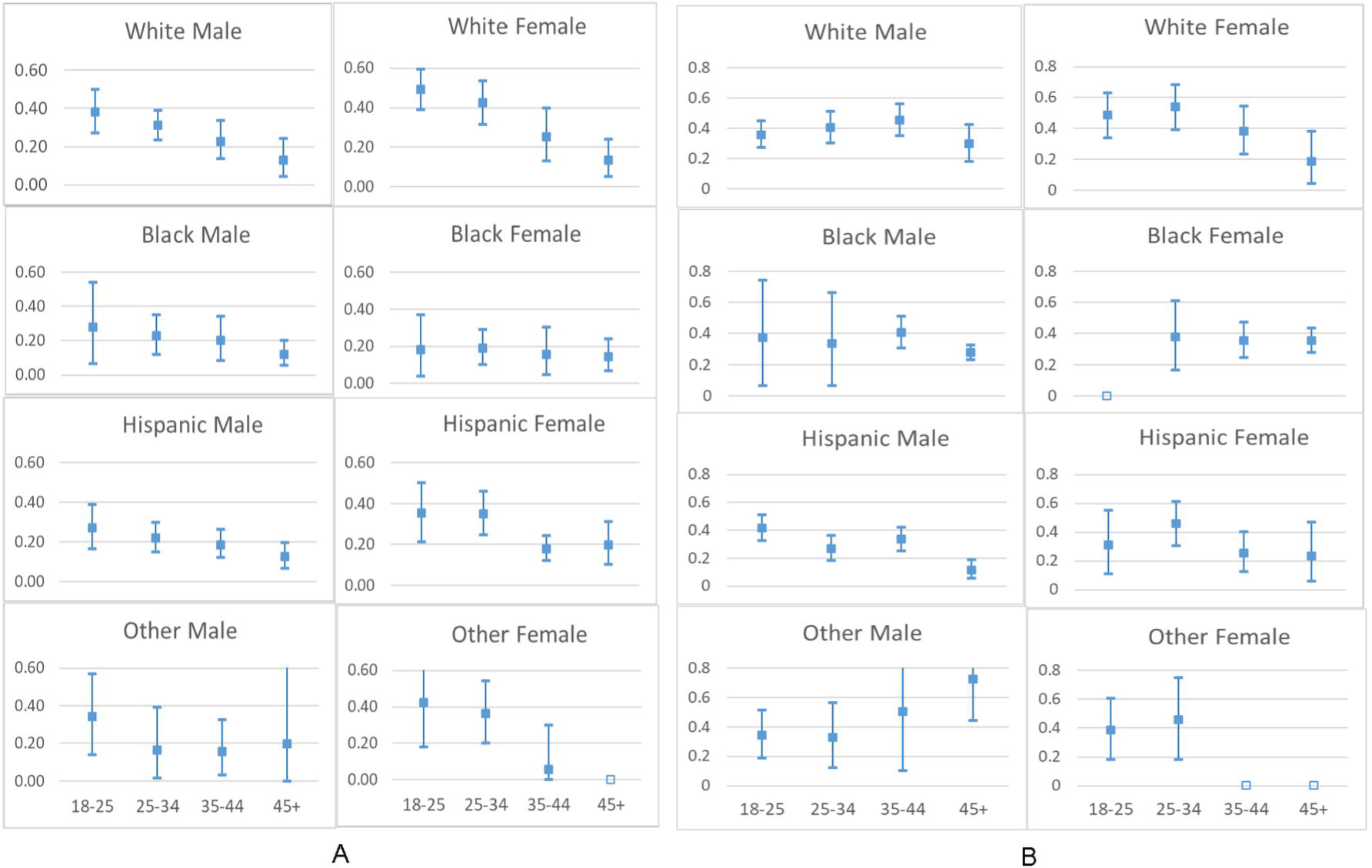
Proportions of PWID reporting (A) receptive syringe sharing. (B) distributive syringe sharing. Filled box indicates point estimate, whiskers indicate 95% confidence interval; empty box indicates insufficient data for estimate.

**Fig 2.**
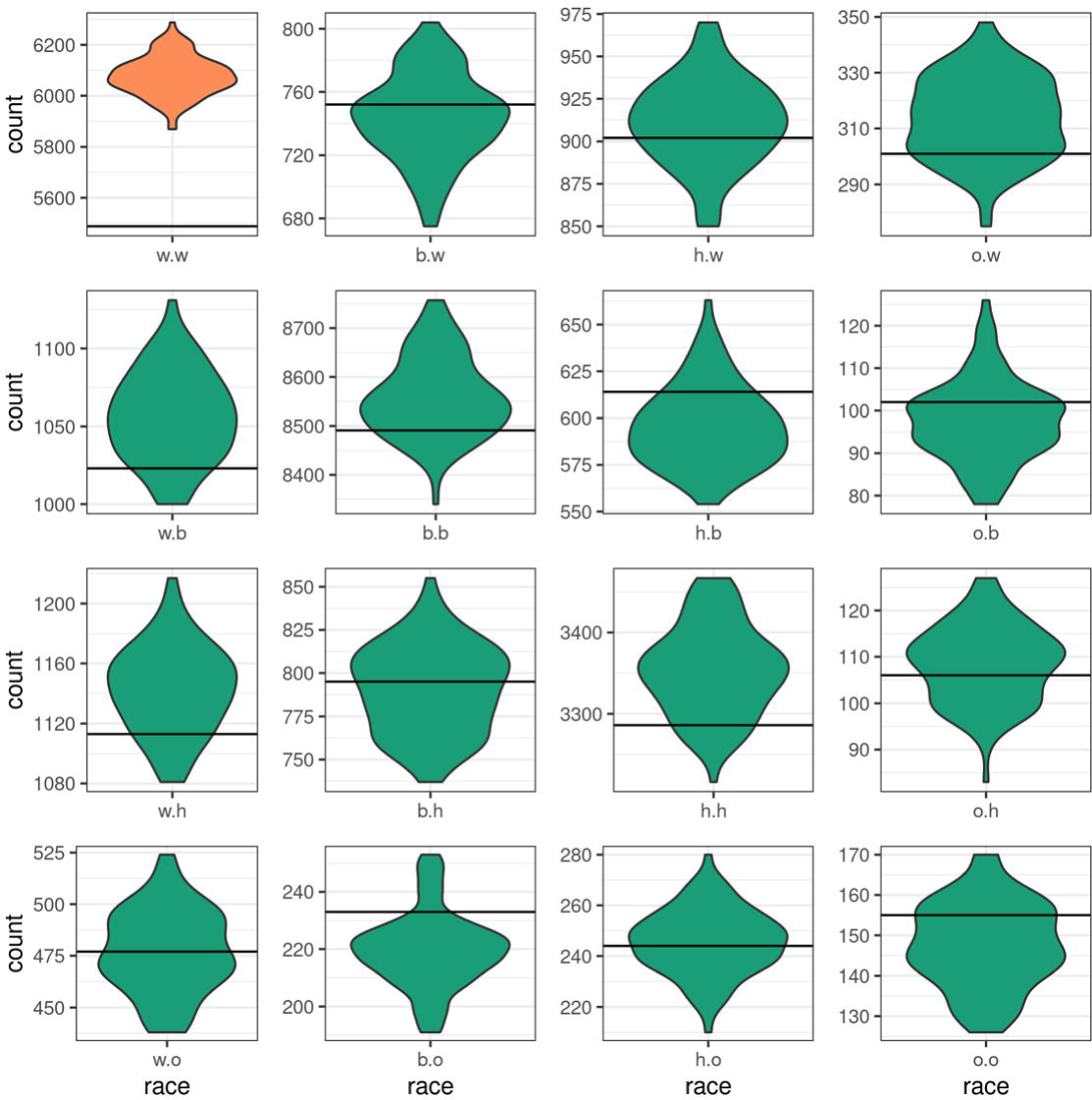
Simulated and target statistics for race mixing. The labels “w”, “b”, “h” and “o” represent White, Black, Hispanic and Other. The White-White term (shown in orange) was left out to avoid collinearity.

In mixed effects regressions, women were more likely than men to report equipment sharing (OR=1.32, 95% CI 1.17-1.50), RSS (OR=1.47, 95% CI 1.37-1.59) and DSS (OR=1.38, 95% CI 1.26-1.52). There were also significant effects of age on all three behaviors. The likelihood of recent equipment sharing decreased with age (Chi2[3]=10.80, p=.01), from 0.69 in the youngest (18-24) category to 0.61 and 0.58 in the intermediate age categories, and 0.56 in the oldest (≥ 45) category. The likelihood of recent RSS also decreased with age (Chi2[3] = 15.18, p=.002), from 0.37 for the 18-24 category to 0.31 and 0.28 in the intermediate categories, and 0.23 for 45 and older. The likelihood of recent DSS was significantly lower for the oldest age category (≥ 45) compared to all other categories (Chi2[1]=31.02, p<.0001), dropping from 0.42 in the youngest category and 0.41 and 0.39 in the intermediate categories, to 0.29 in the oldest category. Hispanics were less likely to report equipment sharing compared to the other race/ethnicity categories (Chi2[1] = 13.5, p=.0002; Hispanic vs. white OR=0.80, 95% CI 0.71-0.90), while RSS was more likely to be reported by white compared to Black or Hispanic PWID (Chi2[3] = 12.47, p=.006; Black vs. white OR = 0.75, 95% CI 0.56-1.01; Hispanic vs. white OR = 0.82, 95% CI 0.70-0.95). The likelihood of DSS was not influenced by race/ethnicity. Persons residing in the city of Chicago were less likely to report RSS (29% vs. 34%, OR = 0.79, 95% CI 0.67 – 0.92) compared to those living outside of Chicago, even while adjusting for other variables.

#### Sharing partners

Among PWID who shared any injection equipment, the estimated average number of sharing partners was 3.5 (95% CI 2.7-4.3). There were no significant effects of demographic variables in negative binomial mixed effects regressions on equipment sharing partners.

The estimates of number of receptive and distributive syringe sharing partners are shown in Tables 4 and 5. Among PWID who used a shared syringe, the overall estimated average number of syringe-sharing partners was 2.3 (95% CI 1.79-2.83). Among PWID who gave a used syringe to another person to use, the estimated average number of people shared with was 2.4 (95% CI 1.84-3.00).

**Table 4.**
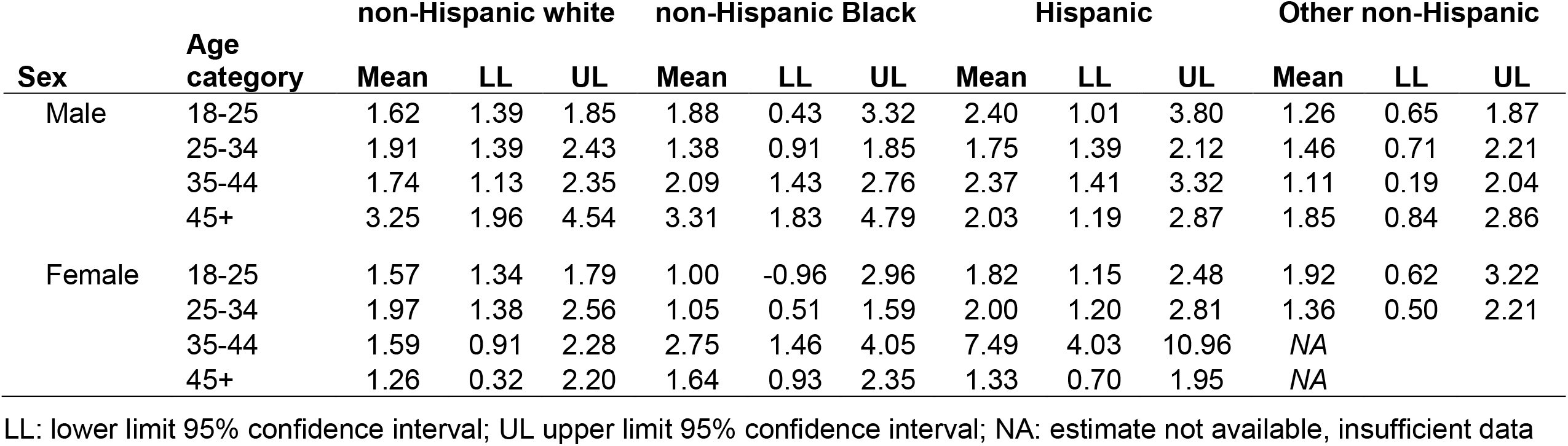
Meta-analysis estimates of number of receptive syringe-sharing (RSS) injection partners among PWID reporting recent RSS, by sex, race or ethnicity, and age category.

**Table 5.** Meta-analysis estimates of number of distributive syringe sharing (DSS) injection partners among PWID reporting recent DSS, by sex, race or ethnicity, and age category.

| Sex | Age category | non-Hispanic white |  |  | non-Hispanic Black |  |  | Hispanic |  |  | Other non-Hispanic |  |  |
| --- | --- | --- | --- | --- | --- | --- | --- | --- | --- | --- | --- | --- | --- |
|  |  | Mean | LL | UL | Mean | LL | UL | Mean | LL | UL | Mean | LL | UL |
| Male | 18-25 | 1.83 | 1.53 | 2.13 | NA |  |  | 2.10 | 1.51 | 2.68 | 1.81 | 1.05 | 2.57 |
|  | 25-34 | 2.62 | 1.71 | 3.54 | 1.90 | 1.15 | 2.65 | 2.12 | 1.64 | 2.61 | 1.97 | 1.05 | 2.88 |
|  | 35-44 | 4.60 | 2.33 | 6.86 | 2.84 | 2.31 | 3.38 | 3.37 | 1.85 | 4.90 | NA |  |  |
|  | 45+ | 3.41 | 1.63 | 5.19 | 3.29 | 2.51 | 4.07 | 2.19 | -0.07 | 4.44 | 2.78 | 1.74 | 3.81 |
| Female | 18-25 | 1.90 | 1.52 | 2.28 | NA |  |  | 2.27 | 1.13 | 3.41 | 1.80 | 0.92 | 2.68 |
|  | 25-34 | 2.80 | 1.76 | 3.85 | 1.22 | 0.46 | 1.99 | 2.50 | 1.37 | 3.63 | 1.83 | 0.57 | 3.10 |
|  | 35-44 | 2.01 | 1.29 | 2.73 | 2.24 | 0.29 | 4.19 | 2.08 | 0.44 | 3.72 | NA |  |  |
|  | 45+ | 2.00 | 0.76 | 3.24 | 2.97 | 1.60 | 4.33 | 2.00 | 0.61 | 3.39 | NA |  |  |

LL: lower limit 95% confidence interval; UL upper limit 95% confidence interval; NA: estimate not available, insufficient data

In mixed effects negative binomial regressions, there were significant effects of age on number of RSS partners (Chi2[3] = 12.92, p=.005) and DSS partners (Chi2[3] = 18.88, p=.0003). The marginal mean numbers of RSS and DSS partners were 2.1 and 2.2, respectively in the youngest age category, and 2.5 and 2.6, respectively in the oldest age category. There was also a significant effect of race/ethnicity on number of DSS partners (Chi2[3] = 27.00, p<.0001), with non-Hispanic other race/ethnicity reporting fewer DSS partners (marginal mean 2.0; vs. white IRR=0.82, 95% CI 0.72-0.94). There was no difference in number of RSS or DSS partners by sex or urban residence.

### Fitting and simulating synthetic syringe sharing networks

#### Model fit

We define the random edge probability as the number of observed edges divided by the maximum number of possible edges on a directed network with 32,000 nodes. We report the ratio (R_p_) of the probability of the edge corresponding to a specified network parameter relative to a random edge. Estimated R_p_ for edges representing needles shared from males to females is 2.41, from females to males is 2.50, and between males is 1.43, indicating the higher probability of syringe sharing from males to females, and between females than between males. The R_p_ values for mixing between age categories are: 5.98 for persons ≤25 years of age; 1.98 for edges representing syringes shared from persons ≤25 years to persons >26 years and 0.74 for edges representing syringe sharing from persons >26 years to persons ≤25 years. We also note that syringe sharing shows strong race homophily as evidenced by the R_p_ values for edges between Black (9.07), Hispanic (5.57), and all other race/ethnicities (9.86) relative to the much lower R_p_ values representing edges between different races (S1 File, Table A1). Syringe sharing between PWID (nodes) residing less than 1/8 mile or 1/8 – 1 mile apart are much more likely (R_p_ values of 1511 and 58.13, respectively); R_p_ for an edge between nodes 1 – 20 miles apart is about 0.8. See S1 File for a detailed summary of the estimated model coefficients and relative probabilities.

#### Comparing the fit of the simulated models to the targets

The distributions of the specified network parameters across the 100 simulated networks and the target values for each of the statistics is shown in Table 6. The target values of each of the network parameters specified in the model (see S1 File) are within the 2.5^th^ and 97.5^th^ percentiles of the simulated distribution. **Table 6. Simulated network parameters and target statistics**

**Table 6.** Simulated network parameters and target statistics.

| Parameter | Target Statistic | Mean (2.5, 97.5 percentiles)<sup>a</sup> |
| --- | --- | --- |
| Number of edges | 24650 | 24840 (24504, 25149) |
| Number of nodes with out-degree |  |  |
| Degree 0 | 20376 | 20315 (20167, 20451) |
| Degree 1 | 5368 | 5382 (5245, 5485) |
| Degree 2 | 2578 | 2590 (2497, 2685) |
| Degree 3 | 1424 | 1429 (1365, 1494) |
| Degree > 3<sup>b</sup> | 2254 | 2284 (2238, 2343) |
| Number of nodes with in-degree: |  |  |
| Degree 0 | 23162 | 23117 (22991, 23264) |
| Degree 1 | 4159 | 4178 (4079, 4281) |
| Degree > 1<sup>b</sup> | 4679 | 4705 (4636, 4773) |
| Race mixing: |  |  |
| Black-White | 752 | 744 (688, 791) |
| Hispanic-White | 902 | 911 (854, 964) |
| Other-White | 301 | 313 (291, 339) |
| White-Black | 1023 | 1056 (1002, 1107) |
| Black-Black | 8491 | 8559 (8438, 8727) |
| Hispanic-Black | 614 | 596 (561, 643) |
| Other-Black | 102 | 98 (80, 119) |
| White-Hispanic | 1113 | 1143 (1085, 1198) |
| Black-Hispanic | 795 | 791 (747, 838) |
| Hispanic-Hispanic | 3286 | 3353 (3259, 3460) |
| Other-Hispanic | 106 | 108 (94, 125) |
| White-Other | 477 | 477 (439, 517) |
| Black-Other | 233 | 219 (193, 250) |
| Hispanic-Other | 244 | 245 (222, 269) |
| Other-Other | 155 | 148 (129, 170) |
| White-White <sup>b</sup> | 5488 | 6077 (5941, 6222) |
| Gender-mixing: |  |  |
| Female-Male | 7487 | 7580 (7463, 7750) |
| Male-Female | 6784 | 6770 (6597, 6926) |
| Male-Male | 7674 | 7694 (7532, 7843) |
| Female-Female <sup>b</sup> | 2699 | 2797 (2715, 2887) |
| Age-mixing |  |  |
| Young-Old | 3170 | 3132 (3037, 3223) |
| Old-Young | 1008 | 990 (935, 1056) |
| Old-Old | 1474 | 1488 (1429, 1556) |
| Young-Young <sup>b</sup> | 18992 | 19230 (18949, 19487) |
| Distance (miles): |  |  |
| Category 1 (< 1/8) | 3870 | 3970 (3948, 3995) |
| Category 2 (1/8 – 1) | 8652 | 8722 (8609, 8835) |
| Category 3 (1 – 20) | 5940 | 5980 (5827, 6131) |
| Category 4<sup>b</sup> (>20) | 5423 | 6168 (5943, 6354) |
| <sup>a</sup> across 100 simulated networks |  |  |
| <sup>b</sup> category omitted for estimation |  |  |

## Discussion

As we would expect, PWID have high rates of joblessness and homelessness. Young white suburban male PWID are somewhat better off than others. This might in part explain why young male PWID are less likely to obtain syringes from a SSP, as they are able to purchase them at drug stores more conveniently. However, this means they are less likely to be exposed to harm reduction services such as syringe exchange, HIV/HCV testing and counseling, and drug and HIV/HCV treatment referrals. Older PWID were both more likely to use a SSP and less likely to report sharing of syringes and other equipment. However, older PWID who did share syringes tended to have more sharing partners than their younger counterparts, which could be an indication of higher HIV and HCV infection risk for this group. In contrast to age effects, the findings for women were inconsistent. Although women were more likely than men to use a SSP, they were also more likely to report sharing of syringes and other equipment. PWID living in Chicago were less likely to report RSS, perhaps reflecting greater access to SSPs. Yet, in spite of greater access to SSPs in Chicago, non-Hispanic Black PWID were less likely than others of the same age to use a SSP. This highlights the need for greater outreach efforts needed to reach Black urban PWID who are at high risk for HCV infection and opioid overdose.

ERGMs allow for fitting of a model that incorporates mixing and degree parameters that describe processes that govern formation of syringe sharing networks in a statistically robust fashion. In this study, we estimated the important impacts of network characteristics such as sex, age, race and ethnicity, and geographic proximity, in addition to the network processes that govern the process of sharing and receiving a syringe (i.e, out- and in-ties, respectively). We found that syringe sharing from males to females and between females was more likely than syringe sharing between males. We also estimated the impact of geographic proximity and race-based homophily on the syringe sharing in the population.

## Conclusions

Computational models are useful in understanding the complex interplay of multilevel factors that might impact the HCV epidemic among PWID. However, these models need to be informed with real socio-behavioral data. Estimates derived from the synthesis of data from multiple studies on a circumscribed population can be used to inform dynamic network models. Future work will incorporate these syringe sharing network parameters in a dynamic simulation model that allows for the generation of a realistic synthetic population and underlying network structures.

## Supporting information

Technical Appendix

## Data Availability

Data are available on Open Science Framework

http://dx.doi.org/10.17605/OSF.IO/4PXZW

## Acknowledgements

The data for this study came from multiple studies funded by the National Institute on Drug Abuse (USA) and the Centers for Disease Control and Prevention (USA). We thank Dr.Lawrence Ouellet and the Chicago Department of Public Health for providing access to several data sets, and we extend our appreciation to Community Outreach Intervention Projects for providing access to service program data.

## Supporting Information

**S1 File. Technical Appendix**. Model fitting procedure, code, and table of estimated coefficients, standard errors and mean edge probability and probability ratio obtained from Exponential Random Graph Model

**S2 File. Figs S1-S3**. Figures showing estimates for proportions of PWID reporting sharing of ancillary injection equipment (Fig S1), syringe-mediated drug sharing (Fig S2), and obtaining syringes from a syringe service program (Fig S3)

