## Supplementary material for "A meta-analysis of 20 years of data on people who inject drugs in metropolitan Chicago to inform computational modeling": Technical Appendix

The theoretical framework for modeling IDU networks is based upon the exponential random graph models (ERGMs),[1] described elsewhere and implemented in the *ergm*[2] packages in the R programming language[3]. Networks were simulated using directed graphs. The log-odds of formation of each partnership type were dependent upon the number and distribution of existing syringe-sharing relationships (henceforth, "relationships") within the network. The mean number of such relationships was estimated from meta-analyses as described above, based on reported numbers of receptive and distributive syringe sharing partners, and the distribution of relationships was determined by other parameters, particularly: mixing based on sex ("male" and "female"), race/ethnicity ("non-Hispanic white", "non-Hispanic Black", "Hispanic", "non-Hispanic other"), and age ("under 25" vs. "25 and older"). Additionally, the distribution of geographic distances across the edges was modeled using a custom-coded ERGM term, and the distribution of in- and out-degree edges was fit as per the meta-mixing data described in the main body of the manuscript. The fitted model is:

$$\text{logit}(\text{p}(y_{ij,t} = 1 | Y^c_{ij,t-1}, y_{ij,t-1} = 0)) = \theta\delta_e e + \sum_{d_0=0}^{3} \theta_{d_o}\delta_{d_o}(\eta(d_o)) + \sum_{d_i=0}^{1} \theta_{d_i}\delta_i(\eta(d_i))$$

$$\sum_{i,j=1}^{2} \theta_{m(a_i,a_j)}\delta_{m(a_i,a_j)}m(a_i, a_{j_i}) + \sum_{i,j=1}^{4} \theta_{m(r_i,r_j)}\delta_{m(r_i,r_j)}m(r_i, r_j) + \sum_{i,j=1}^{4} \theta_{m(g_i,g_j)}\delta_{m(g_i,g_j)}m(g_i, g_j)$$

(A.1)

where $e$ is the number of edges; $\eta(d_o)$ is the distribution of outdegrees $d_o = \{0, 1, 2, 3\}$; in-degrees $d_i = \{0, 1\}$; $m(a_i, a_j)$is the number of ties between individuals in age category 1 and age category 2; $m(r_i, r_j)$ is the number of ties between edges between individuals in each of the four age categories; $m(g_i, g_j)$ is the number of ties between edges between individuals in each of the two gender categories. The functions corresponding to each of these model terms represent the change in their value corresponding to the "toggle" of one dyad (defined as removing one existing tie, or adding a non-existent one); the change statistic functions are needed to estimate the coefficients using Markov Chain Monte Carlo techniques, as per the algorithmic routines in *ergm*. MCMC convergence was assessed using diagnostic convergence tests (not shown).

The model incorporates a new parameter *dist* that is not included in the standard library of ERGM packages to compute the set of distances across which all edges in the network are distributed. The parameter *dist* is coded using the techniques provided in Hunter, Goodreau, Handcock (henceforth referred to as HGH)[4]. Four geographic distance categories are defined, as explained in the main body of the manuscript, using location information for each agent, with location coded using the agent's longitude and latitude. The distance between any two nodes as computed here uses the equirectangular approximation to determine straight-line distances between two points on earth.

The core computation in fitting an ERGM with the *dist* term requires the computation of the "change statistic", intuitively understood as the difference in a network statistic obtained by "toggling" one particular edge (ie, adding an edge where none exists or removing an existent edge). The distribution of edges in the four distance categories are jointly independent and hence the *dist* term coded here is a dyadic independent term (full technical details on the computation of change statistics and supporting references are available in HGH). Coding a parameter that allows for estimation of the change in log-odds associated with toggling a particular edge in the incorporates a "wrapper function" to be called by the user written in R and a function to compute the change statistic in C. The change in the network statistic for dist associated with the toggle of one edge can be computed locally without full information of the rest of the network.

**Figure A.1: Wrapper function in R.**

| Code | Explanation |
|---|---|
| `InitErgmTerm.dist <- function(nw, arglist, ...) {` | As per HGH |
| ` a <- check.ErgmTerm(nw, arglist, directed=NULL,` |  |
| `bipartite=FALSE,` | As per HGH |
| ` varnames = c("dist"),` | Define ERGM parameter `dist` |
| ` vartypes = c("numeric"),` | Specify type numeric |
| ` required = c(TRUE),` | As per HGH |
| ` defaultvalues = list(NULL))` | As per HGH |
| ` dist<-a$dist` | Extract covariate from network object |
| ` if(length(dist)==0){return(NULL)}` | Specification error check |
| ` coef.names <- paste("dist",dist,sep="")` | As per HGH |
| ` name <- "dist"` | As per HGH |
| ` nodelat <- get.node.attr(nw, "lat")` | Input needed to compute distance: latitude |
| ` nodelon <- get.node.attr(nw, "lon")` | Input needed to compute distance: longitude |
| ` list(name = name,` | As per HGH |
| ` coef.names = coef.names,` |  |
| ` pkgname = "ergm.userterms",` |  |
| ` inputs = c(dist, nodelat, nodelon),` |  |
| ` dependence = FALSE` |  |
| ` )` |  |
| `}` | |

### C-side

**Figure A.2: Computation of the change statistic: C**

| Code | Explanation |
|---|---|
| `#include "changestats.users.h"` | As per HGH |
| `#include <math.h>` | Needed header |
| `#ifndef M_PI` | Define constant |
| ` #define M_PI 3.1415926535` |  |
| `#endif` | |
| `CHANGESTAT_FN(d_dist) {` | |
| ` Vertex t, h;` | Define variables |
| ` int i, j;` |  |
| ` double t_nodecov, h_nodecov;` |  |
| ` int dist_cat, target_cat;` |  |
| ` int change;` |  |
| ` double t_lat, t_lon, h_lat, h_lon;` |  |

```
  double t_lat_rad, t_lon_rad, h_lat_rad, h_lon_rad;
  double radius, xunit, yunit, dist;

  ZERO_ALL_CHANGESTATS(i);
  FOR_EACH_TOGGLE(i) {
    t = TAIL(i); h = HEAD(i);
    t_lat = INPUT_PARAM[t + N_CHANGE_STATS - 1];
    t_lon = INPUT_PARAM[t + N_NODES + N_CHANGE_STATS - 1];
    h_lat = INPUT_PARAM[h + N_CHANGE_STATS - 1];
    h_lon = INPUT_PARAM[h + N_NODES + N_CHANGE_STATS - 1];
    //printf("%f,%f,%f,%f\n", t_lat,t_lon,h_lat,h_lon);
    // Turn degrees into radians
    t_lat_rad = M_PI*t_lat/180.0;
    t_lon_rad = M_PI*t_lon/180.0;
    h_lat_rad = M_PI*h_lat/180.0;
    h_lon_rad = M_PI*h_lon/180.0;

// Average radius of the earth (kilometers)
    radius = 6334.02;

    // Equirectangular approximation (good for close
distances)
    xunit = (h_lon_rad - t_lon_rad) * cos((t_lat_rad +
h_lat_rad)/2);
    yunit = h_lat_rad - t_lat_rad;
    dist = sqrt( (xunit * xunit) + (yunit * yunit) ) *
radius;

    // Find dist category
    //printf("%f\n", dist);
    if (dist < 0.2){
      target_cat = 1;
    } else if (dist < 1.6) {
      target_cat = 2;
    } else if (dist < 32.2) {
      target_cat = 3;
    } else target_cat = 4;

    change = IS_OUTEDGE(t,h) ? -1 : 1;
    for(j = 0; j < N_CHANGE_STATS; j++) {
      if ((int)INPUT_PARAM[j] == target_cat){
        CHANGE_STAT[j] += change;
```

Method for extracting tail and head node attributes

Constant needed for equirectangular approximation

Converting longitude and latitude to x and y coordinates and computation of distance

Check to see if the toggled edge was removed or added

```
      }
    }
    TOGGLE_IF_MORE_TO_COME(i);
  }
  UNDO_PREVIOUS_TOGGLES(i);
}
```

The parameters estimated from (1) are given below.

**Table A.1: Estimated coefficients, standard errors and mean edge probability and probability ratio obtained from Exponential Random Graph Model given in Equation (1).**

| Parameter | Estimated coefficient | Standard Error | p-value | Edge probability | Probability Relative to Random Edge ($R_p$)** |
|---|---|---|---|---|---|
| Edges* | -10.13 | 0.04 | 1.00E-04 | 3.99E-05 | 1.66 |
| mix.gender.male.female† | 0.37 | 0.02 | 1.00E-04 | 5.81E-05 | 2.41 |
| mix.gender.female.male | 0.41 | 0.02 | 1.00E-04 | 6.02E-05 | 2.50 |
| mix.gender.male.male | -0.15 | 0.02 | 1.00E-04 | 3.44E-05 | 1.43 |
| mix.young.1.0‡ | 0.13 | 0.02 | 1.00E-04 | 4.56E-05 | 1.89 |
| mix.young.0.1 | -0.81 | 0.03 | 1.00E-04 | 1.77E-05 | 0.74 |
| mix.young.1.1 | 1.28 | 0.02 | 1.00E-04 | 1.44E-04 | 5.98 |
| mix.race.num.2.1§ | -1.03 | 0.04 | 1.00E-04 | 1.42E-05 | 0.59 |
| mix.race.num.3.1 | -0.81 | 0.04 | 1.00E-04 | 1.78E-05 | 0.74 |
| mix.race.num.4.1 | -0.08 | 0.06 | 1.63E-01 | 3.68E-05 | 1.53 |
| mix.race.num.1.2 | -1.39 | 0.03 | 1.00E-04 | 9.99E-06 | 0.41 |
| mix.race.num.2.2 | 1.70 | 0.01 | 1.00E-04 | 2.18E-04 | 9.07 |
| mix.race.num.3.2 | -0.88 | 0.04 | 1.00E-04 | 1.65E-05 | 0.69 |
| mix.race.num.4.2 | -0.82 | 0.10 | 1.00E-04 | 1.75E-05 | 0.73 |
| mix.race.num.1.3 | -0.95 | 0.03 | 1.00E-04 | 1.54E-05 | 0.64 |
| mix.race.num.2.3 | -0.30 | 0.04 | 1.00E-04 | 2.95E-05 | 1.22 |
| mix.race.num.3.3 | 1.21 | 0.02 | 1.00E-04 | 1.34E-04 | 5.57 |
| mix.race.num.4.3 | -0.39 | 0.10 | 1.00E-04 | 2.69E-05 | 1.12 |
| mix.race.num.1.4 | -0.10 | 0.04 | 2.47E-02 | 3.63E-05 | 1.51 |
| mix.race.num.2.4 | 0.18 | 0.06 | 3.75E-03 | 4.79E-05 | 1.99 |
| mix.race.num.3.4 | 0.31 | 0.06 | 1.00E-04 | 5.47E-05 | 2.27 |
| mix.race.num.4.4 | 1.78 | 0.08 | 1.00E-04 | 2.37E-04 | 9.86 |
| idegree0 | 3.96 | 0.04 | 1.00E-04 | 2.09E-03 | 86.79 |
| idegree1 | 1.57 | 0.03 | 1.00E-04 | 1.91E-04 | 7.95 |
| odegree0 | -3.28 | 0.13 | 1.00E-04 | 1.50E-06 | 0.06 |

| | | | | | |
|---|---|---|---|---|---|
| odegree1 | -3.39 | 0.10 | 1.00E-04 | 1.35E-06 | 0.06 |
| odegree2 | -2.57 | 0.08 | 1.00E-04 | 3.05E-06 | 0.13 |
| odegree3 | -1.62 | 0.05 | 1.00E-04 | 7.92E-06 | 0.33 |
| dist1 (< 1/8 mile) | 6.85 | 0.02 | 1.00E-04 | 3.64E-02 | 1511.22 |
| dist2 (1/8 – 1 mile) | 3.56 | 0.02 | 1.00E-04 | 1.40E-03 | 58.13 |
| dist3 (1 mile – 20 miles) | -0.69 | 0.02 | 1.00E-04 | 2.00E-05 | 0.83 |

*The edges term represents an edge between two white females under age 25 at a distance >20 miles (category 4) from each other, who do not have an in-degree of 0 or 1 or an outdegree of 0, 1, 2, or 3.
**Ratio of edge probability relative to a random edge between any two nodes. The random edge probability is computed as the number of observed edges divided by the maximum number of possible edges on a directed network with, in this case, 32,000 nodes.
† Suffix .male.female represents syringe given from male to female.
‡ The term mix.young.1.0 represents syringes given from young.1 (≤ 25 years of age) to young.0 (>26 years of age)
§ Race categories: race.num.1 = White; race.num.2 = Black; race.num.3 = Hispanic; race.num.4 = Other. mix.race.num.2.1 represents syringes given from Black to White persons.

One cell in each of the mixing matrices and the distance 4 category were left unspecified to avoid model collinearity (see Morris et al.[5] for details on overspecification of ERGMs). Specified models converged up to indegrees sssss0 and 1 and outdegrees 0, 1, 2, and 3; hence the in- and out-degree terms were specified to these values.

The log-odds corresponding to the edge term are estimated to be -10.12, which, per our fitted model, represents an edge between two white females under age 25 at a distance >20 miles (category 4) from each other, who do not have an in-degree of 0 or 1 or an outdegree of 0, 1, 2, or 3 (henceforth, “base edge”). The probability of a base edge is exp(-10.12)/exp(1+-10.12) = 3.99 e-05. (For comparison, *any* edge in this model has estimated log odds of -10.63, corresponding to a probability of 2.41 e-05.) The probability of an edge relative to a random edge is denoted by $R_{p.}$ A base edge in the model is about 1.66 times as likely as a random edge (i.e., $R_p$=1.66). Estimated coefficients for specific parameters represent the change in the log-odds corresponding to that parameter. For instance, the log odds of an edge between two males is 0.15 units lower than the log odds of the base edge; in probability terms, the $R_p$ for a male-to-male edge is 1.43. The interpretation of other edge terms in the model is similar. Edges that result in a unit increase in the number of nodes with indegree 1 or a unit decrease in edges with in-degree 0 have $R_p$ values of about 8 and 87. The model convergence was assessed using the MCMC diagnostics as recommended in the *statnet* package.

One hundred networks were simulated from model (1). The target statistics for each of the fitted parameters was within the IQR of the distribution across the 100 simulated networks. The distribution for the race mixing term is given in the main body of the manuscript and remaining figures are given here.

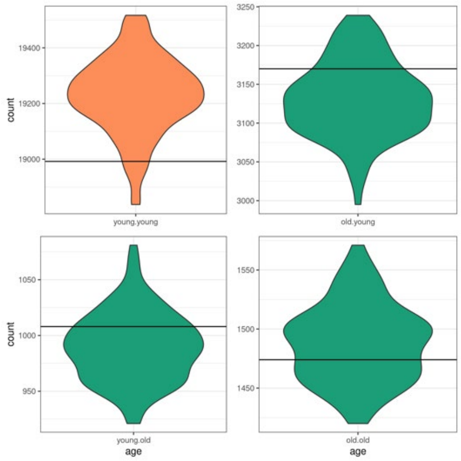

(A) Age mixing

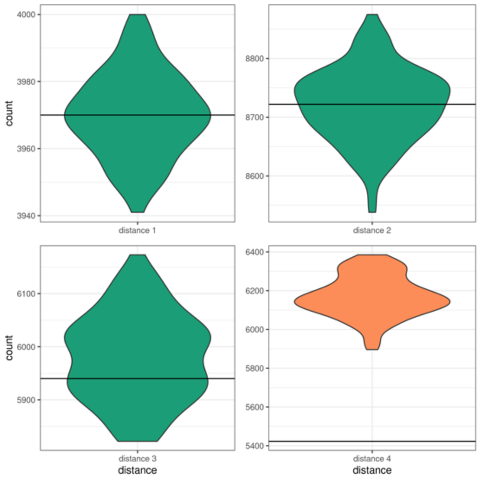

(B) Distance

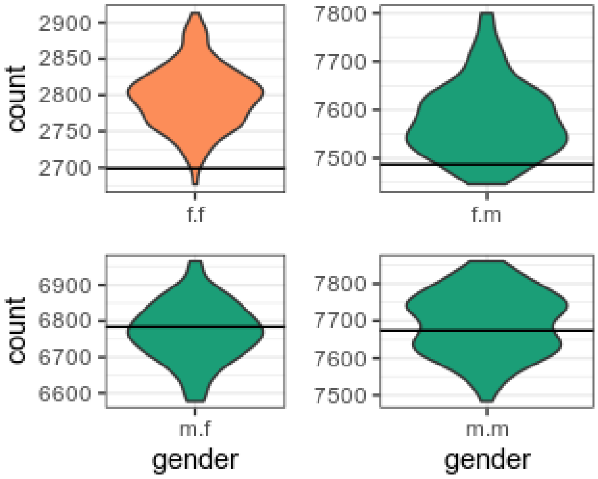

C: Sex mixing

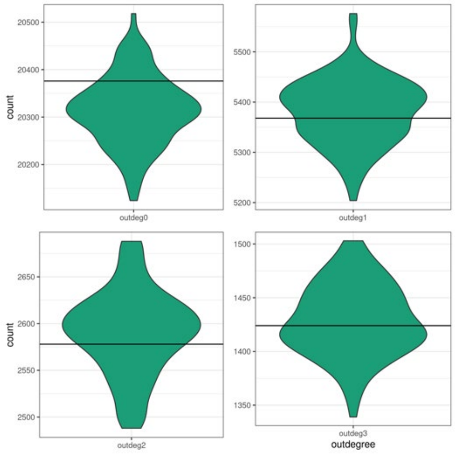

(D) Outdegrees

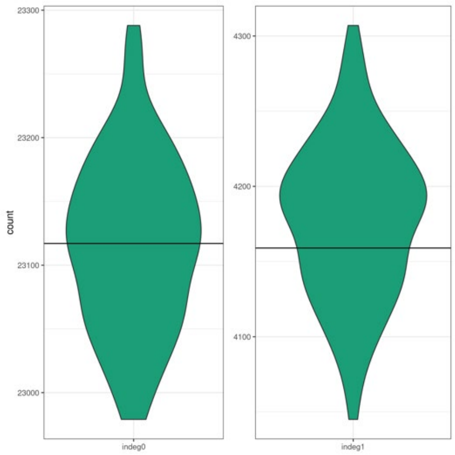

(E) Indegrees

Figure A.3: Target statistics (solid line) and simulated interquartile ranges for parameters in the model (1). Specified Model terms are in green. Unspecified model terms are in orange.
