## Supplementary material for "A meta-analysis of 20 years of data on people who inject drugs in metropolitan Chicago to inform computational modeling": Technical Appendix

*Supplemental Figures*

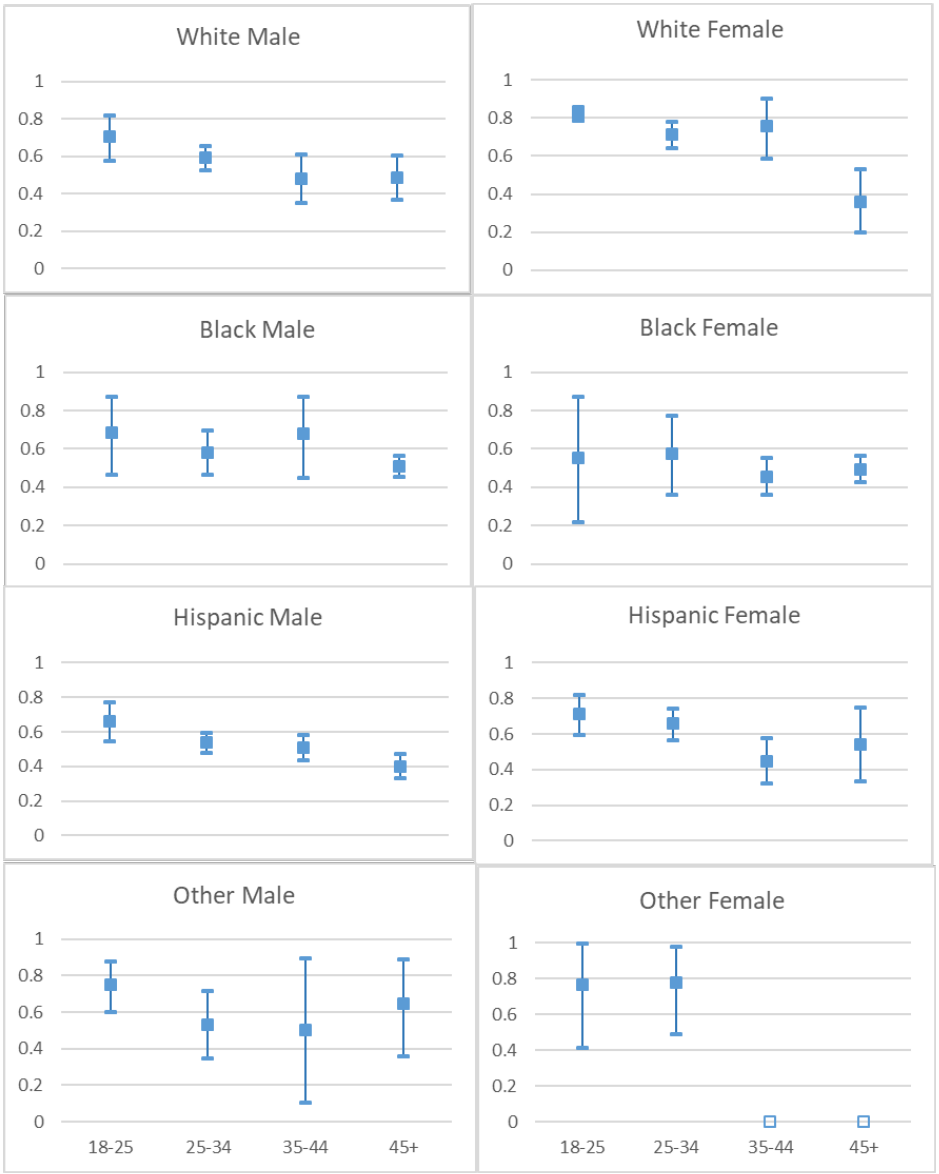


*Fig S1. Proportions of PWID reporting equipment sharing*

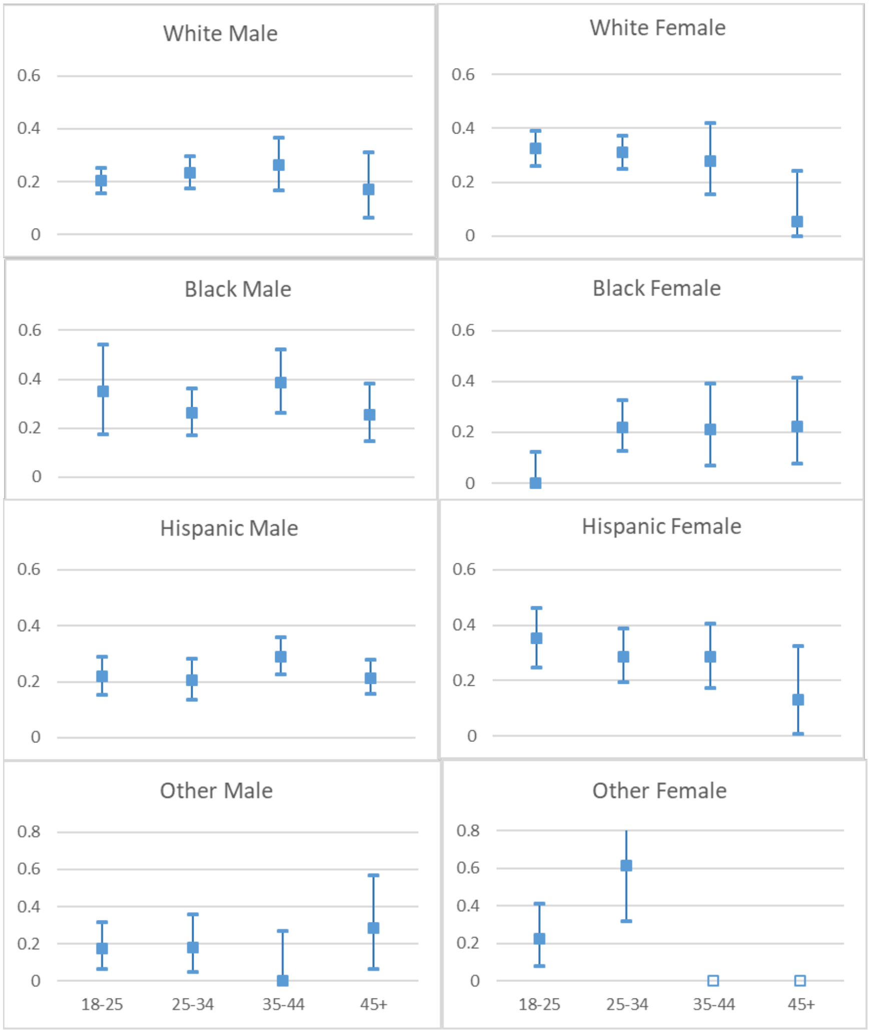


*Fig S2. Proportions of PWID reporting syringe mediated drug sharing*

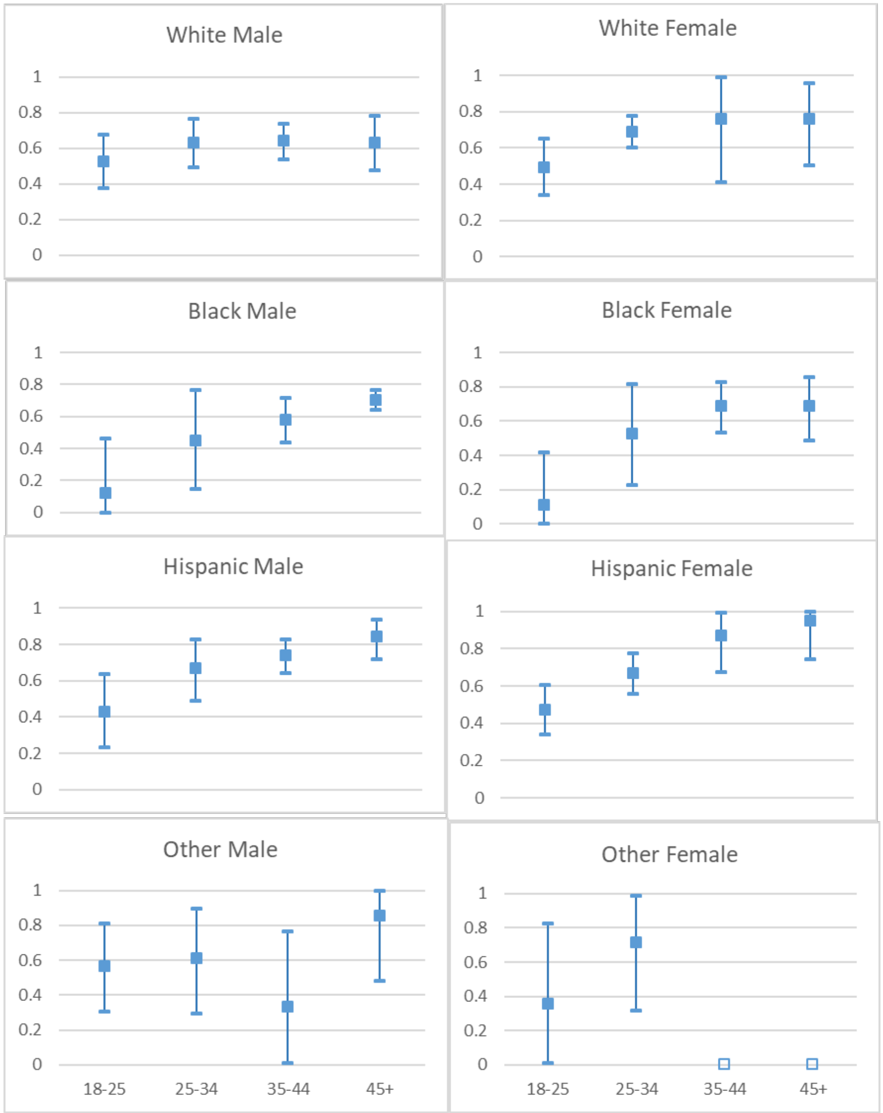


*Fig S3. Obtained syringes from a syringe exchange program*
